# Upper airway gene expression reveals a more robust innate and adaptive immune response to SARS-CoV-2 in children compared with older adults

**DOI:** 10.1101/2021.07.15.21260285

**Authors:** Eran Mick, Alexandra Tsitsiklis, Natasha Spottiswoode, Saharai Caldera, Paula Hayakawa Serpa, Angela M. Detweiler, Norma Neff, Angela Oliveira Pisco, Lucy M. Li, Hanna Retallack, Kalani Ratnasiri, Kayla M. Williamson, Victoria Soesanto, Eric A. F. Simões, Amy Kistler, Brandie D. Wagner, Joseph L. DeRisi, Lilliam Ambroggio, Peter M. Mourani, Charles R. Langelier

**Author notes:** These authors contributed equally (Eran Mick, Alexandra Tsitsiklis). These authors jointly supervised this work (Peter M. Mourani, Charles R. Langelier).

## Abstract

Unlike other respiratory viruses, SARS-CoV-2 disproportionately causes severe disease in older adults and only rarely in children. To investigate whether differences in the upper airway immune response could contribute to this disparity, we compared nasopharyngeal gene expression in 83 children (<19-years-old; 38 with SARS-CoV-2, 11 with other respiratory viruses, 34 with no virus) and 154 adults (>40-years-old; 45 with SARS-CoV-2, 28 with other respiratory viruses, 81 with no virus). Expression of interferon-stimulated genes (ISGs) was robustly activated in both children and adults with SARS-CoV-2 compared to the respective non-viral groups, with only relatively subtle distinctions. Children, however, demonstrated markedly greater upregulation of pathways related to B cell and T cell activation and proinflammatory cytokine signaling, including TNF, IFNγ, IL-2 and IL-4 production. Cell type deconvolution confirmed greater recruitment of B cells, and to a lesser degree macrophages, to the upper airway of children. Only children exhibited a decrease in proportions of ciliated cells, the primary target of SARS-CoV-2, upon infection with the virus. These findings demonstrate that children elicit a more robust innate and adaptive immune response to SARS-CoV-2 infection in the upper airway that likely contributes to their protection from severe disease in the lower airway.

## Introduction

One of the defining features of the COVID-19 pandemic has been the striking relationship between disease severity and age^1–3^. While infection with other respiratory viruses, such as influenza or respiratory syncytial virus (RSV), causes significant morbidity and mortality in both young children and older adults^3–9^, severe COVID-19 occurs disproportionately in older adults and only very rarely in children^1,3,10–14^. A recent comprehensive modeling study estimated that the infection fatality rate is lowest for children ages 5-9 (∼0.001%) and that even adults in their 40s may already be at 100-fold greater risk of death from COVID-19^1^. The age-dependent effect on disease severity and mortality has been shown even when accounting for age-associated comorbidities^15^.

A few studies have examined differences in systemic immunological profiles of hospitalized children and adults with COVID-19, revealing greater breadth and neutralizing activity of SARS-CoV-2 specific antibodies as well as stronger CD4+ T cell responses to viral spike protein in adults^16,17^. These studies suggested that poorer outcomes in adults are not due to failure to engage a systemic adaptive immune response, and that children may exhibit a less pronounced adaptive response because their infection course is milder. One study reported higher serum levels of IL-17A and interferon-γ (IFNγ) in children early in their hospitalization^17^, suggesting a more robust innate immune response may contribute to the milder disease course.

Our understanding of age-related differences in the immune response at the site of initial infection, the upper airway, remains limited. Several large-scale studies have found no systematic differences between children and adults in the distribution of SARS-CoV-2 viral load measured in nasopharyngeal (NP) swabs^18,19^, suggesting children are not generally better able to control viral replication in the upper airway. However, differences in the upper airway microenvironment and immune response could still contribute to protection from severe disease in children, for example, by limiting migration of the virus into the lower airway.

Small-scale studies have begun to compare the upper airway immune response to SARS-CoV-2 in children and adults, with some contradictory results. One study found that children with COVID-19 expressed higher levels of genes associated with innate immune pathways, including interferon-stimulated genes and genes related to NLRP3 inflammasome signaling^20^. Another study, however, found no age-related differences in interferon-stimulated gene expression and reported globally similar host transcriptional responses between adults and children with diverse types of viral infections^21^, highlighting the need for further investigation. Importantly, neither study directly controlled for SARS-CoV-2 viral load when comparing immune-related gene expression between children and adults.

We previously used metagenomic RNA-sequencing of NP swabs to compare upper airway gene expression in adult patients with COVID-19, other viral acute respiratory illnesses or non-viral illnesses^22^. Our analysis revealed a pronounced interferon response in COVID-19 patients, proportional to SARS-CoV-2 viral load, but attenuated activation of additional innate immune and pro-inflammatory pathways compared to patients with other viral infections^22^. Here, we report new sequencing data from a relatively large pediatric cohort to enable assessment of age-related differences in the upper airway transcriptional response to SARS-CoV-2 infection and shed further light on these outstanding questions. When controlling for viral load, our results suggest that differences in the overall magnitude of interferon-stimulated gene expression in the upper airway of children and adults with COVID-19 are relatively subtle and seem unlikely to explain their distinct clinical outcomes. However, we also find clear evidence of more robust pro-inflammatory and adaptive immune responses in the upper airway of children, which may contribute to their protection from severe disease.

## Results and Discussion

To compare the upper airway gene expression response to SARS-CoV-2 infection in children and adults, we utilized our previously published dataset of NP swab RNA-sequencing from an adult cohort alongside newly sequenced swabs from a pediatric cohort. All samples in both cohorts were obtained in the course of clinical testing for SARS-CoV-2 by reverse transcription polymerase chain reaction (RT-PCR) at the University of California San Francisco or Children’s Hospital Colorado. We included patients up to 19 years of age in the pediatric cohort and restricted the adult cohort to those 40 years of age or older to impose clearer age separation.

As in our previous analysis, we divided each age cohort into three viral status groups: 1) patients with PCR-confirmed SARS-CoV-2 infection (“SARS-CoV-2” group), 2) patients negative for SARS-CoV-2 by PCR with no other pathogenic respiratory virus detected by metagenomic RNA sequencing (“No Virus” group), and 3) patients negative for SARS-CoV-2 who had another respiratory virus detected by sequencing (“Other Virus” group). Finally, we limited the samples in the SARS-CoV-2 group to those with at least 10 viral reads-per-million (rpM), comparable to PCR C_t_ values below 30^22^, which have been associated with recovery of actively replicating virus from respiratory specimens^23–25^.

The final dataset included 83 children (38 SARS-CoV-2, 34 No Virus, 11 Other Virus; median age 4, IQR 2-12) and 154 adults (45 SARS-CoV-2, 81 No Virus, 28 Other Virus; median age 62, IQR 47-71) (**Figure 1A,B; Supplemental Table 1A-C**). There were no statistically significant differences between children and adults with respect to the site of clinical testing (**Supplemental Table 1A**). Most of the patients in the SARS-CoV-2 group in both age cohorts were tested as outpatients, indicative of an early/mild stage of disease (**Supplemental Table 1A**). Samples in the SARS-CoV-2 group in both age cohorts spanned several orders of magnitude of viral load, and while viral load trended higher in the children this did not reach statistical significance (**Figure 1C**). Rhinovirus was the most prevalent other respiratory virus detected in both age cohorts and a diversity of other viruses were also identified (**Figure 1D**).

**Figure 1:**
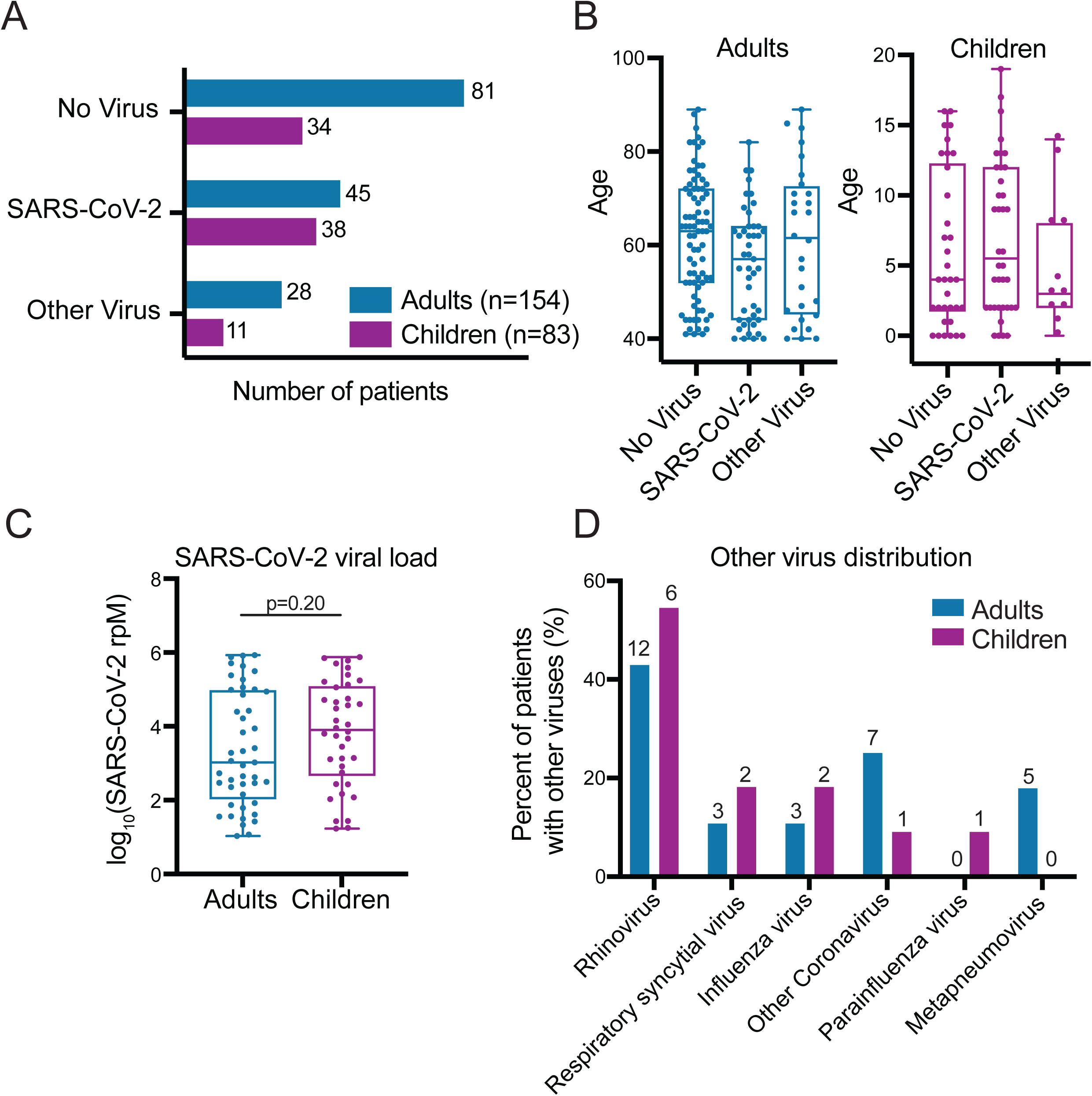
Cohort characteristics. **A)** Number of patients in the SARS-CoV-2, No Virus and Other Virus groups in the adult and pediatric cohorts. **B)** Age distribution across the three viral status groups in the adult and pediatric cohorts. **C)** Distribution of SARS-CoV-2 viral load, measured in reads-per-million (rpM), in adult and pediatric SARS-CoV-2 patients. P-value derives from a two-sided Mann-Whitney test. **D)** Distribution of viruses in the Other Virus groups in the adult and pediatric cohorts. Absolute numbers are provided above each bar while the y-axis indicates percentage out of each cohort’s Other Virus group. One child had both influenza and rhinovirus and two adults had both RSV and rhinovirus.

We began by performing differential expression (DE) analyses between the SARS-CoV-2 and No Virus groups within each age cohort separately. This approach minimizes confounding by age-related gene expression differences unrelated to SARS-CoV-2 infection and any potential batch effects, though it could be affected by differences among the patients in each cohort’s No Virus group. The analyses yielded 1,961 and 1,216 differentially expressed genes at an adjusted p-value < 0.1 for the pediatric and adult cohorts, respectively (**Supplemental Data 1**). We then performed gene set enrichment analyses^26^ (GSEA) using Gene Ontology (GO) biological process annotations^27^ on the DE results in each cohort and compared the enriched pathways.

As expected, a range of immune related pathways were upregulated in both adults and children with SARS-CoV-2 compared to those with no virus (**Figure 2A; Supplemental Data 2**). Pathways related to the interferon response appeared as a whole to be strongly induced in both children and adults. Children, however, demonstrated stronger upregulation of the B cell and T cell activation pathways, macrophage activation and phagocytosis, and several cytokine production pathways, including IL-2, IL-4, TNF and IFNγ. A few immune pathways showed inconsistent expression changes compared to the respective No Virus groups, including neutrophil mediated immunity and mast cell mediated immunity. Intriguingly, both children and adults with SARS-CoV-2 infection exhibited downregulation of olfactory receptor gene expression (sensory perception of chemical stimulus pathway), consistent with the loss of sense of smell clinically observed in both age cohorts^28,29^.

**Figure 2:**
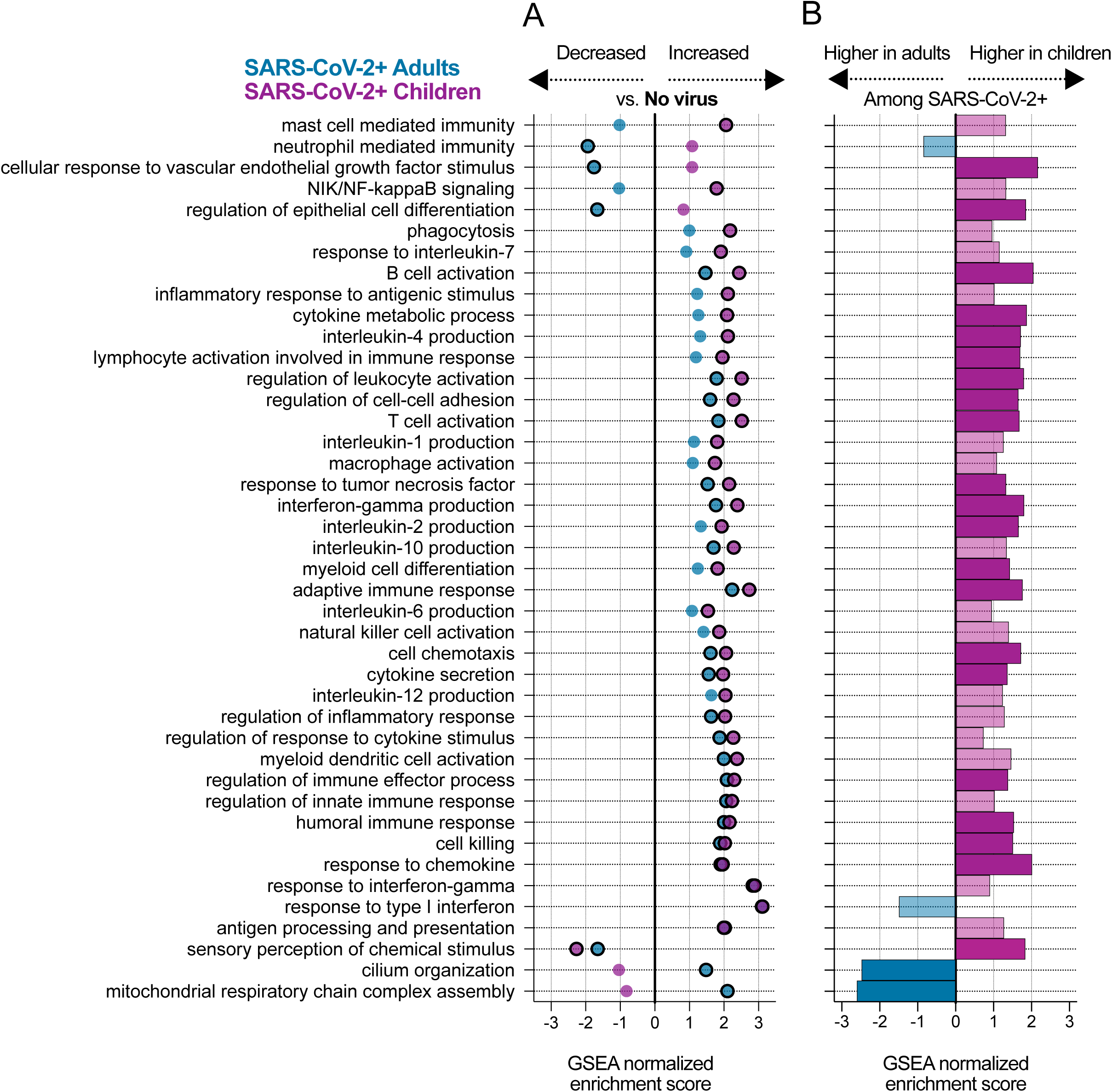
Pathways activated in children and adults upon SARS-CoV-2 infection. **A)** Normalized enrichment scores of selected GO biological process terms that reached statistical significance (Benjamini-Hochberg adjusted P-value < 0.05) in the GSEA using DE genes between the SARS-CoV-2 and No Virus groups in either the adult or pediatric cohort. Statistical significance is denoted by a black outline around the circle. **B)** Normalized enrichment scores for the same GO terms as in (A) in the GSEA using DE genes between children and adults with SARS-CoV-2. Dark color bars represent pathways that reached statistical significance (Benjamini-Hochberg adjusted P-value < 0.05).

We complemented the analyses comparing SARS-CoV-2 and No Virus patients in each cohort separately by directly comparing gene expression between SARS-CoV-2 infected children and adults, controlling for viral load. We identified 5,352 differentially expressed genes at an adjusted p-value < 0.1 (**Supplemental Data 3**), and GSEA of the DE results yielded overall similar patterns to those described above (**Figure 2B; Supplemental Data 4**). In particular, B cell related pathways (B cell activation, humoral immune response), T cell related pathways (T cell activation, IFNγ production, IL-2 and IL-4 production) and chemokine/cytokine signaling were more highly expressed in children with COVID-19.

While some immune pathways did not reach statistical significance in the direct comparison between children and adults with SARS-CoV-2, they typically trended in the same direction (**Figure 2B**). On the other hand, the stark disparity in neutrophil activation observed in the comparison to the No Virus groups was only weakly supported in the direct comparison, likely reflecting differences among the No Virus patients themselves. The direct comparison clearly revealed lower expression of cilia-associated genes in children with SARS-CoV-2 and also suggested a trend toward lower expression of interferon-stimulated genes, though the pathway just missed the statistical significance cutoff. As expected, many developmental processes unrelated to infection also differed in the direct comparison between children and adults (**Supplemental Data 4**).

Many of the pathways identified in the GSEA results as differentially expressed between children and adults with COVID-19 were tightly related to particular cell types. We therefore applied *in silico* estimation of cell type proportions^30^ based on marker genes derived from an airway single-cell study^31^ as an additional approach to contextualize our findings (**Figure 3, Figure 3–figure supplement 1; Supplemental Data 5**).

Consistent with the GSEA results, we found that SARS-CoV-2 infection triggered significantly greater recruitment of B cells to the upper airway in children compared to adults, which was also evident in the comparison between children and adults with other respiratory viruses (**Figure 3A**). In contrast, differences in estimated T cell proportions were much subtler (**Figure 3–figure supplement 1**), suggesting the GSEA results reflected distinctions in T cell identity and regulation rather than cell number.

**Figure 3:**
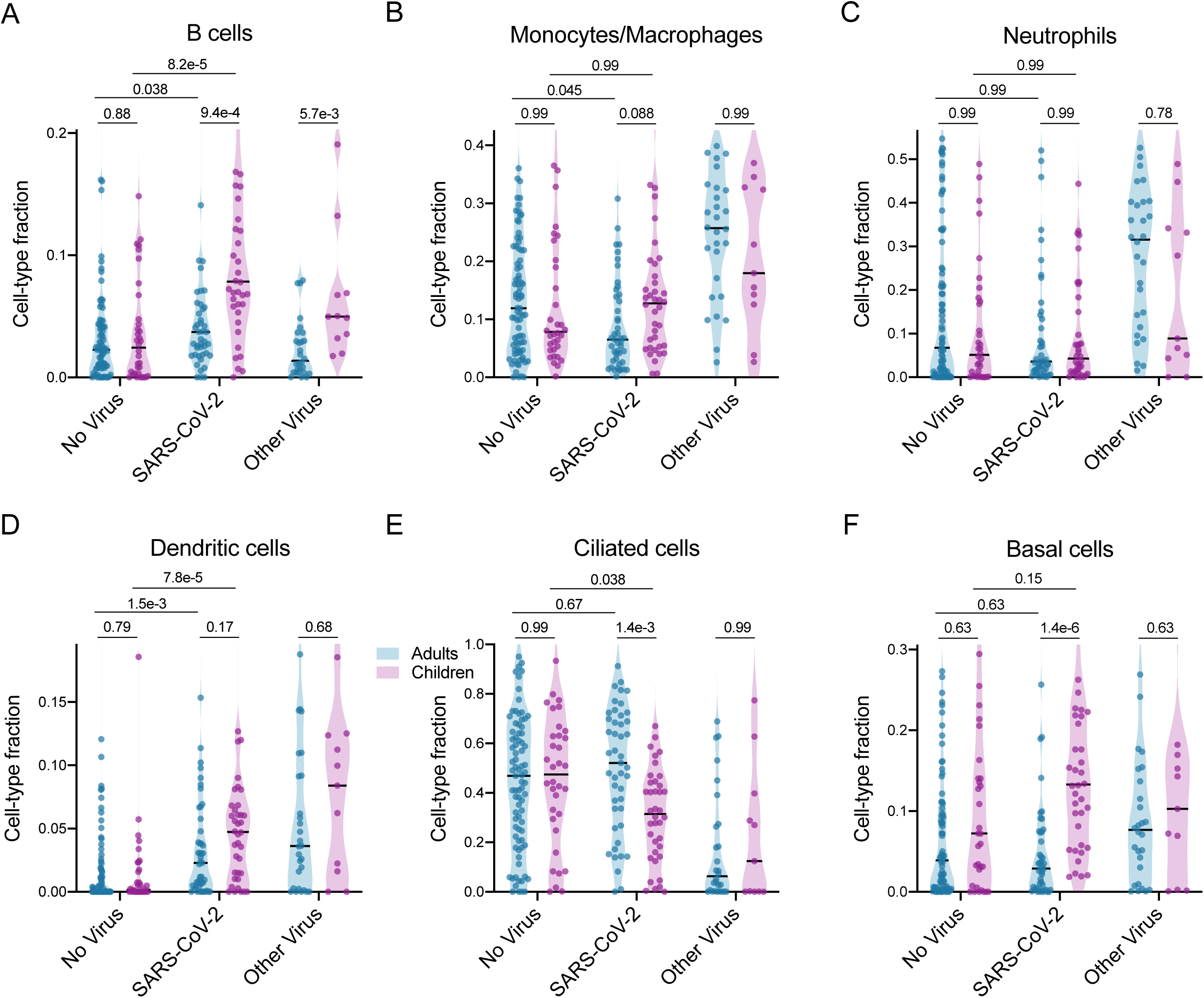
Cell-type proportion differences between children and adults. *In silico* estimation of cell-type proportions in the bulk RNA-sequencing using single-cell signatures. Black lines denote the median. The y-axis in each panel was trimmed at the maximum value among all groups of 1.5*IQR above the third quartile, where IQR is the interquartile range. For each cell type, we formally compared each viral status group between the two age cohorts as well as the No Virus and SARS-CoV-2 groups within each age cohort. Pairwise comparisons were performed with a two-sided Mann-Whitney test followed by Holm’s correction for multiple testing.

We previously observed in our adult study that infection with SARS-CoV-2 was associated with blunted recruitment of macrophages and neutrophils to the upper airway as compared to other respiratory viruses^22^, and these trends appear to re-capitulate in children despite the limited sample size of the pediatric Other Virus group (**Figure 3B,C**). Nevertheless, macrophage proportions trended higher in children with SARS-CoV-2 compared to adults (**Figure 3B**), as did those of dendritic cells (**Figure 3D**).

Intriguingly, while proportions of ciliated cells did not differ between children and adults in the No Virus group, children with SARS-CoV-2 exhibited a marked decrease in ciliated cell proportions that was entirely absent in the adults (**Figure 3E**), consistent with the GSEA findings. This was accompanied by greater proportions of basal cells in children compared to adults with SARS-CoV-2 (**Figure 3F**). Recent studies have found that ciliated cells are a major target for SARS-CoV-2 at the onset of infection^32,33^, and a possible interpretation of our results is that children are better able to turn over and clear infected ciliated cells in the nasopharynx, potentially helping them more effectively control the infection.

Finally, we examined the effect of SARS-CoV-2 viral load on gene expression in pathways of interest within each age cohort using robust regression. Expression of interferon-stimulated genes (ISGs) frequently correlates with viral load, as we observed in our previous analysis in adults^22^. Children and adults exhibited similar trends for several of the ISGs that most strongly correlated with viral load in our previous analysis, such as *CXCL11* and *OASL*, though the slope trended higher in children (**Figure 4A**). However, some of the ISGs that were strongly differentially expressed in adults with COVID-19 compared to those with non-viral illnesses exhibited much more gradual activation in the pediatric cohort, such that only children with high viral load achieved comparable expression to that of adults (e.g., *IFI6, IFI27*; **Figure 4B**). The “lagging” response of these ISGs in children with lower SARS-CoV-2 viral load explains the trend toward lower pathway expression observed in the GSEA results (**Figure 2B**). These findings suggest differences in ISG-specific regulation and/or cellular origins between children and adults that defy simple generalization. While the significance of such subtle distinctions is unclear, they appear less likely to drive the dramatic outcome disparities.

**Figure 4:**
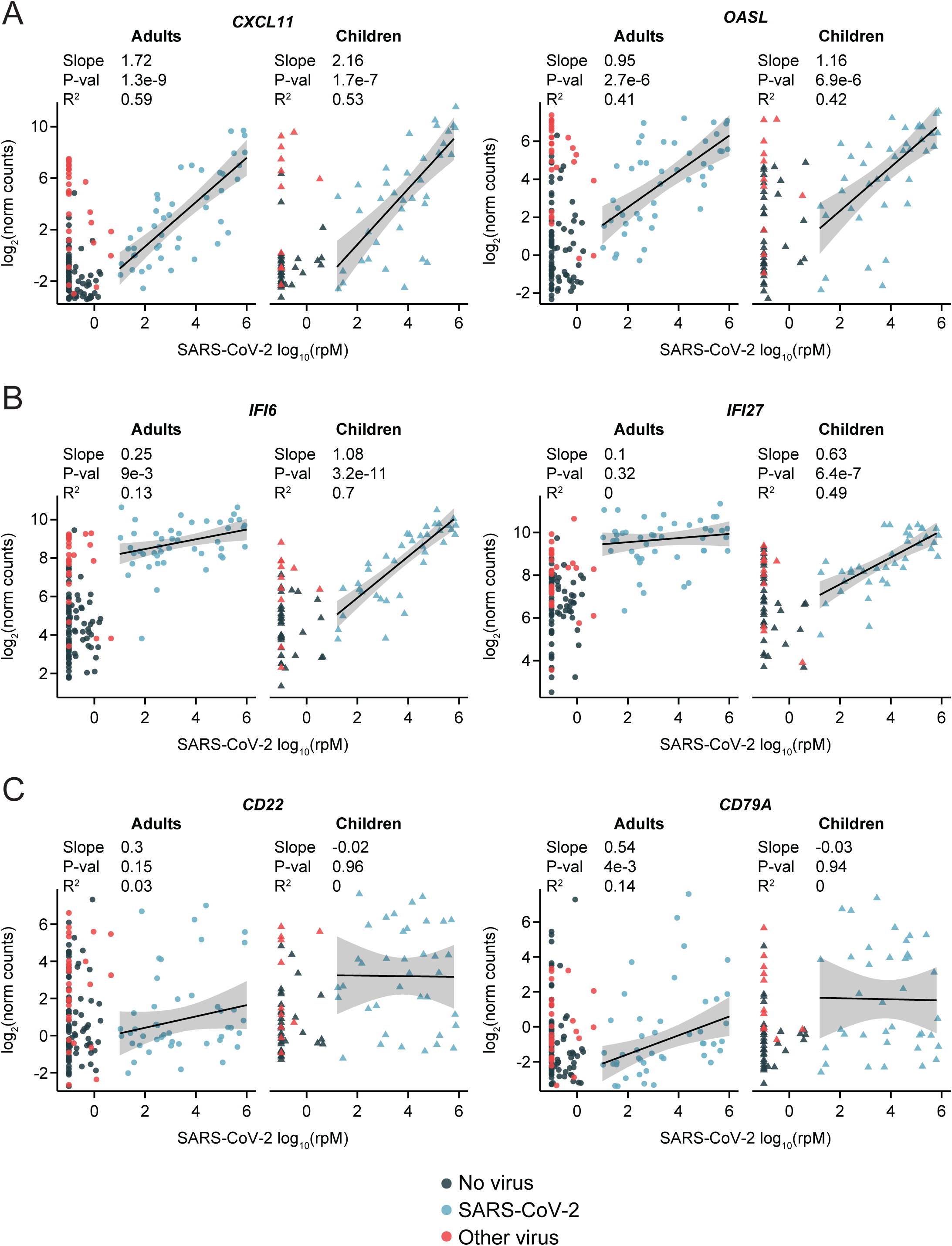
Relationship of selected immune pathway marker genes to SARS-CoV-2 viral load in children and adults. Scatter plots of normalized gene counts (log_2_ scale, y-axis) as a function of SARS-CoV-2 viral load (log_10_(rpM), x-axis) in each age cohort. The viral status group is indicated by the dot color. Robust regression was performed on SARS-CoV-2 patients to characterize the relationship to viral load. Shaded bands represent 95% confidence intervals. Numerical results listed for each gene refer to, from top to bottom: the regression slope, the p-value for the difference of the slope from 0, and the adjusted robust coefficient of determination (R^2^). **A)** Canonical type I interferon response genes showing high correlation to viral load in adults and children. **B)** Interferon-stimulated genes showing a more gradual response to viral load in children compared to adults. **C)** B cell marker genes.

In stark contrast to ISGs, the expression of several B cell marker genes (e.g. *CD22, CD79A)* was weakly correlated with viral load, especially in children (**Figure 4C**). These genes exhibited significant heterogeneity between patients, likely reflecting the timing of activation of the B cell response, but the fraction of children who were engaging the response at the time of sampling was clearly greater.

Our study has several limitations that should be kept in mind: 1) a larger sample size would have increased the generalizability of our findings; 2) we could not directly control for timing since onset of infection; 3) proteomic measurements may have provided additional insight but were not possible from the available swab specimens, which were collected into a virus inactivating agent; 4) our findings regarding tissue cellular composition require validation by single-cell sequencing; and lastly, 5) the majority of subjects with COVID-19 had mild disease at the time of sampling and did not require hospitalization. Results may have differed if specimens from a greater proportion of severely ill individuals had been available, however, this likely resulted in a more relevant comparison since few children develop severe disease.

Taken together, our findings suggest children elicit a more robust pro-inflammatory and adaptive immune response to SARS-CoV-2 infection in the upper airway compared to adults but an overall comparable type I interferon response. Children, unlike adults, also display decreased proportions of ciliated cells, the primary targets of SARS-CoV-2, upon infection with the virus. These differences in the upper airway immune response and tissue composition may both contribute to the protection of children from severe disease in the lower respiratory tract. Further study is warranted to unravel why children are better protected specifically against SARS-CoV-2, or perhaps more broadly against β-coronaviruses^34^, as compared to other common respiratory viruses.

## Materials and Methods

### Study design and clinical cohort

We previously conducted and published an observational cohort study of adult patients with acute respiratory illnesses tested for COVID-19 by RT-PCR at the University of California, San Francisco (UCSF), leveraging leftover RNA extracted from clinical NP swab specimens^22^. The UCSF Institutional Review Board (IRB) granted a waiver of consent under IRB protocol #17-24056. For the novel analyses presented here, we leveraged the published adult dataset while supplementing it with pediatric samples similarly obtained at UCSF under the same protocol. Additionally, pediatric samples were obtained from patients tested for COVID-19 by RT-PCR from NP swabs at Children’s Hospital Colorado (CHCO). CHCO specimens and data were obtained under IRB protocols #0865, #20-1617 and #20-0972, which also granted a waiver of consent. Demographic and clinical data for all patients were obtained from a combination of electronic and manual abstraction of medical records at the respective institutions.

Children up to 19 years of age and adults at least 40 years of age were eligible for inclusion in the present analysis. Enrollment was not restricted based on sex or race. Some samples were ultimately excluded based on sequencing metrics, as described in the following sections.

### Sample processing

Excess clinical swab specimens were stored in viral transport media at −80°C in the Clinical Microbiology Laboratories. Specimens were thawed and 200uL aliquots of specimen were added to 200uL of DNA/RNA Shield (Zymo Research, Irvine, CA) in sterile 1.5 mL microtubes with appropriate biohazard precautions.

### Metagenomic RNA sequencing

Specimens underwent RNA extraction and metagenomic sequencing, as previously described^22^. Briefly, RNA was extracted from 200μL of specimen in DNA/RNA shield using bead-based lysis and the Zymo Pathogen Magbead kit (Zymo). We also processed negative control samples (water and HeLa cell RNA) to account for background contamination. All samples were spiked with RNA standards from the External RNA Controls Consortium (ERCC)^35^. Depletion of cytosolic and mitochondrial rRNA was performed using Fastselect (Qiagen, Germantown, MD). RNA was reverse transcribed to generate cDNA and used to construct sequencing libraries using the NEBNext Ultra II Library Prep Kit (New England Biolabs, Ipswich, MA). Libraries underwent 146 nucleotide paired-end sequencing on an Illumina Novaseq 6000 instrument.

### Metagenomic analysis of respiratory viruses

Samples were processed through the IDSeq pipeline^36,37^, which performs reference based alignment at both the nucleotide and amino acid level against sequences in the National Center for Biotechnology Information (NCBI) nucleotide (NT) and non-redundant (NR) databases, respectively, followed by assembly of the reads matching each taxon detected. We further processed the results for viruses with established pathogenicity in the respiratory tract^38^. We evaluated whether one of these viruses was present in a patient sample if it met the following three initial criteria: (i) at least 10 counts mapped to NT sequences, (ii) at least 1 count mapped to NR sequences, (iii) average assembly nucleotide alignment length of at least 70 bp.

Negative control (water and HeLa cell RNA) samples enabled estimation of the number of background reads expected for each virus, which were normalized by input mass as determined by the ratio of sample reads to spike-in ERCC RNA standards. Viruses were then additionally tested for whether the number of sequencing reads aligned to them in the NT database was significantly greater than background. This was done by modeling the number of background reads as a negative binomial distribution, with mean and dispersion fitted on the negative controls. We estimated the mean parameter of the negative binomial for each taxon (virus) by averaging the read counts across all negative controls after normalizing by ERCCs. We estimated a single dispersion parameter across all taxa using the functions glm.nb() and theta.md() from the R package MASS. We considered a sample to have a pathogenic respiratory virus detected by sequencing if the virus achieved an adjusted p-value < 0.05 after Holm’s correction for all tests performed in the same sample.

We used the IDSeq-calculated viral reads-per-million (rpM), based on the NT alignment, as a uniform measure of SARS-CoV-2 abundance across all samples. A value of 0.1 rpM was added to all samples with rpM < 0.1.

### Assignment of samples to comparator groups

As in our previous analysis^22^, we divided the samples in each age cohort into three viral status groups: 1) samples with a positive clinical PCR test for SARS-CoV-2 were assigned to the “SARS-CoV-2” group; 2) samples with a negative PCR test for SARS-CoV-2 and no evidence of another pathogenic respiratory virus in the metagenomic sequencing were assigned to the “No Virus” group; and 3) samples with a negative PCR test for SARS-CoV-2 but another respiratory virus detected by sequencing were assigned to the “Other Virus” group.

We retained for analysis only samples in the SARS-CoV-2 groups with at least 10 rpM, roughly corresponding to PCR C_t_ values below 30^22^, to focus on cases with likely active viral replication, where a clear transcriptional response to the virus is expected to be found. This approach was based on the well-established correlation between viral load and recovery of actively replicating virus from respiratory specimens, and more specifically the finding that specimens with a PCR C_t_ < 30 are associated with the ability to culture SARS-CoV-2^23–25^.

### Human gene expression quantification

Following demultiplexing, sequencing reads were pseudo-aligned with kallisto^39^ (v. 0.46.1; including bias correction) to an index consisting of all transcripts associated with human protein coding genes (ENSEMBL v.99), cytosolic and mitochondrial ribosomal RNA sequences, and the sequences of ERCC RNA standards. Samples were retained for analysis if they had at least 400,000 estimated counts associated with transcripts of protein coding genes. Gene-level counts (n=19,939) were generated from the transcript-level abundance estimates using the R package tximport^40^, with the scaledTPM method.

### Differential expression (DE) analyses

Genes were retained for each DE analysis if they had at least 10 counts in at least 20% of the samples included in the analysis. All analyses were performed with the R package limma^41^, using quantile normalization and the voom method. The design formula for the comparisons within each age cohort was ∼viral status, where viral status was either “SARS-CoV-2” or “No Virus”. The design formula for the direct comparison between children and adults with SARS-CoV-2 was ∼log_10_(rpM) + age cohort, where age cohort was either “children” or “adults”.

### Gene set enrichment analyses (GSEA)

Gene set enrichment analysis was based on Gene Ontology (GO) biological process pathway annotations^27^, using the non-redundant version available through WebGestalt^42^. Only pathways with a minimum size of 10 genes and a maximum size of 1,500 genes were retained for analysis. The analysis was performed using the fgseaMultilevel function in the R package fgsea^43^, which calculates p-values based on an adaptive, multilevel splitting Monte Carlo scheme. The input consisted of all genes in the respective DE analysis, pre-ranked by fold-change. The gene sets shown in **Figure 2** were manually selected to reduce redundancy and highlight diverse biological functions from among those with a Benjamini-Hochberg adjusted p-value < 0.05 in at least one of the analyses. Full results are provided in **Supplemental Data 2** and **Supplemental Data 4**.

### *In silico* estimation of cell type proportions

Cell-type proportions were estimated using the CIBERSORT X algorithm^30^ based on single cell signatures derived from the human lung cell atlas^31^. Differences in estimated proportions between comparator groups were evaluated for statistical significance using a Mann-Whitney test with Holm’s correction for multiple testing.

### Regression of gene counts against viral load

We performed robust regression of the limma-generated quantile normalized gene counts (log_2_ scale) against log_10_(rpM) of SARS-CoV-2 for selected genes. The analysis was performed within each age cohort separately using the R package robustbase^44^, which implements MM-type estimators for linear regression^45,46^, the KS2014 setting and the model: quantile normalized counts (log_2_ scale) ∼ log_10_(rpM). Model predictions were generated using the R package ggeffects and used for display in the individual gene plots. Error bands represent normal distribution 95% confidence intervals around each prediction. Reported p-values for significance of the difference of the regression coefficient from 0 are based on a t-statistic. Reported R^2^ values represent the adjusted robust coefficient of determination^47^.

## Data Availability

Gene counts and sample metadata have been deposited under NCBI GEO accession GSE179277. The published human lung single-cell datasets used for cell-type proportions analysis can be obtained through Synapse under accessions syn21560510 and syn21560511.

## Funding

NHLBI K23HL138461-01A1 (CL); Chan Zuckerberg Biohub (AD, AOP, NN, JLD); COVID-Child Health Research Award by the Research Institute at Children’s Hospital Colorado; philanthropic support from Mark and Carrie Casey, Julia and Kevin Hartz, Carl Kawaja and Wendy Holcombe, Eric Keisman and Linda Nevin, Martin and Leesa Romo, Diana Wagner, Jerry Yang and Akiko Yamazaki, and Three Sisters Foundation.

## Acknowledgements

We would like to thank Sam Dominguez, MD, PhD, Kirk Harris, PhD, Aline Maddux, MD, Christina Osborne, MD, and Matthew Leroue, MD, for their input on study design and review of the data.

## Supplemental Tables

**Supplemental Table 1A:** Adult vs Pediatric cohort characteristics.

|  | Adults<br>(All) | Children<br>(All) | *p* value | Adults<br>(SARS-CoV-2) | Children<br>(SARS-CoV-2) | *p* value |
| --- | --- | --- | --- | --- | --- | --- |
| Total Enrolled (n) | 154 | 83 |  | 45 | 38 |  |
| Age, years (median, range, IQR) | 62 (40-89,<br>47-71) | 4 (<1-19, 2-12) |  | 57 (40-82, 44-64) | 5 (<1-19, 2-11) |  |
| Female gender | 78 (51%) | 42 (51%) | 0.93 | 25 (56%) | 16 (42%) | 0.27 |
| **Clinical Encounter Type** |  |  |  |  |  |  |
| Inpatient | 42 (27%) | 31 (37%) |  | 4 (9%) | 1 (3%) |  |
| Intensive Care Unit | 16 (10%) | 0 (0%) |  | 2 (4%) | 0 (0%) |  |
| Emergency Department | 24 (16%) | 19 (23%) |  | 3 (7%) | 6 (16%) |  |
| Outpatient | 55 (36%) | 31 (37%) |  | 24 (53%) | 30 (79%) |  |
| Unknown | 17 (11%) | 2 (2%) | 0.59 | 12 (27%) | 1 (3%) | 0.23 |
| **Race** |  |  |  |  |  |  |
| White or Caucasian | 67 (44%) | 34 (41%) |  | 6 (13%) | 13 (34%) |  |
| Asian | 25 (16%) | 6 (7%) |  | 6 (13%) | 1 (3%) |  |
| Black or African American | 14 (9%) | 4 (5%) |  | 1 (2%) | 2 (5%) |  |
| Native Hawaiian or Pacific Islander | 1 (1%) | 2 (2%) |  | 1 (2%) | 0 (0%) |  |
| American Indian or Alaska Native | 0 (0%) | 1 (1%) |  | 0 (0%) | 0 (0%) |  |
| Other | 27 (18%) | 29 (35%) |  | 17 (38%) | 17 (45%) |  |
| Unknown | 20 (13%) | 7 (8%) | **0.013** | 14 (31%) | 5 (13%) | 0.10 |
| **Ethnicity** |  |  |  |  |  |  |
| Not Hispanic or Latino | 109 (71%) | 43 (52%) |  | 16 (36%) | 10 (26%) |  |
| Hispanic or Latino | 23 (15%) | 34 (41%) |  | 14 (31%) | 23 (61%) |  |
| Unknown | 22 (14%) | 6 (7%) | **<0.001** | 15 (33%) | 5 (13%) | 0.06 |

Values are n (%) unless otherwise indicated.
\*Race categories with <1 patient in both groups were excluded from analyses.
Unknown values were excluded from analyses.

**Supplemental Table 1B:** Adult cohort clinical and demographic characteristics.

|  | Cohort Overall | SARS-CoV-2 | Other Virus | No Virus | p value |
| --- | --- | --- | --- | --- | --- |
| Total Enrolled (n) | 154 | 45 | 28 | 81 |  |
| Age, years (median, range, IQR) | 62 (40-89, 47-71) | 57 (40-82, 44-64) | 61.5 (40-89, 45-73) | 62 (41-89, 52-72) | 0.75 |
| Female gender | 78 (51%) | 25 (56%) | 10 (36%) | 43 (53%) | 0.21 |
| **Clinical Encounter Type** |  |  |  |  |  |
| Inpatient | 42 (27%) | 4 (9%) | 9 (32%) | 29 (36%) |  |
| Intensive Care Unit | 16 (10%) | 2 (4%) | 5 (18%) | 9 (11%) |  |
| Emergency Department | 24 (16%) | 3 (7%) | 6 (21%) | 15 (19%) |  |
| Outpatient | 55 (36%) | 24 (53%) | 8 (29%) | 23 (28%) |  |
| Unknown | 17 (11%) | 12 (27%) | 0 (0%) | 5 (6%) | **0.003** |
| **Race** |  |  |  |  |  |
| White or Caucasian | 67 (44%) | 6 (13%) | 19 (68%) | 42 (52%) |  |
| Asian | 25 (16%) | 6 (13%) | 5 (18%) | 14 (17%) |  |
| Black or African American | 14 (9%) | 1 (2%) | 1 (4%) | 12 (15%) |  |
| Native Hawaiian or Pacific Islander | 1 (1%) | 1 (2%) | 0 (0%) | 0 (0%) |  |
| American Indian or Alaska Native | 0 (0%) | 0 (0%) | 0 (0%) | 0 (0%) |  |
| Other | 27 (18%) | 17 (38%) | 3 (11%) | 7 (9%) |  |
| Unknown | 20 (13%) | 14 (31%) | 0 (0%) | 6 (7%) | **<0.001\*** |
| **Ethnicity** |  |  |  |  |  |
| Not Hispanic or Latino | 109 (71%) | 16 (36%) | 26 (93%) | 67 (83%) |  |
| Hispanic or Latino | 23 (15%) | 14 (31%) | 1 (4%) | 8 (10%) |  |
| Unknown | 22 (14%) | 15 (33%) | 1 (4%) | 6 (7%) | **<0.001** |

Values are n (%) unless otherwise indicated.
\*patients with Native Hawaiian or Pacific Islander race; and patients with American Indian or Alaska Native race were excluded from statistical analysis due to insufficient numbers.
Unknown values were excluded from analyses.
Age was analyzed as one-way ANOVA and all categorical variables using chi-squared tests.

**Supplemental Table 1C:** Pediatric cohort clinical and demographic characteristics.

|  | Cohort Overall | SARS-CoV-2 | Other Virus | No Virus | p value |
| --- | --- | --- | --- | --- | --- |
| Total Enrolled (n) | 83 | 38 | 11 | 34 |  |
| Age, years (median, range, IQR) | 4 (<1-19, 2-12) | 5 (<1-19, 2-11) | 3 (<1-14, 2-11) | 4 (<1-16, 2-12) | 0.97 |
| Female gender | 42 (51%) | 16 (42%) | 7 (64%) | 19 (56%) | 0.38 |
| **Clinical Encounter Type** |  |  |  |  |  |
| Inpatient | 31 (37%) | 1 (3%) | 2 (18%) | 28 (82%) |  |
| Intensive Care Unit | 0 (0%) | 0 (0%) | 0 (0%) | 0 (0%) |  |
| Emergency Department | 19 (23%) | 6 (16%) | 8 (73%) | 5 (15%) |  |
| Outpatient | 31 (37%) | 30 (79%) | 0 (0%) | 1 (3%) |  |
| Unknown | 2 (2%) | 1 (3%) | 1 (9%) | 0 (0%) | **<0.001** |
| **Race** |  |  |  |  |  |
| White or Caucasian | 34 (41%) | 13 (34%) | 4 (36%) | 17 (50%) |  |
| Asian | 6 (7%) | 1 (3%) | 1 (9%) | 4 (12%) |  |
| Black or African American | 4 (5%) | 2 (5%) | 2 (18%) | 0 (0%) |  |
| Native Hawaiian or Pacific Islander | 2 (2%) | 0 (0%) | 1 (9%) | 1 (3%) |  |
| American Indian or Alaska Native | 1 (1%) | 0 (0%) | 0 (0%) | 1 (3%) |  |
| Other | 29 (35%) | 17 (45%) | 2 (18%) | 10 (29%) |  |
| Unknown | 7 (8%) | 5 (13%) | 1 (9%) | 1 (3%) | 0.31** |
| **Ethnicity** |  |  |  |  |  |
| Not Hispanic or Latino | 43 (52%) | 10 (26%) | 7 (64%) | 26 (76%) |  |
| Hispanic or Latino | 34 (41%) | 23 (61%) | 3 (27%) | 8 (24%) |  |
| Unknown | 6 (7%) | 5 (7%) | 1 (9%) | 0 (0%) | **<0.001** |

Values are n (%) unless otherwise indicated.
\*No pediatric patients were admitted to the intensive care unit; this category was excluded from statistical analyses.
\*\*patients with Native Hawaiian or Pacific Islander race; patients with American Indian or Alaska Native race; and patients with Black or African American race were excluded from statistical analysis due to insufficient numbers.
All unknown values were excluded from analyses.
Age was analyzed as one-way ANOVA and all categorical variables using chi-squared tests.

## Supplemental Data

**Supplemental Data 1**. Differential expression results between the SARS-CoV-2 and No Virus groups in adults and children separately.

**Supplemental Data 2**. GSEA applied to the differential expression results between the SARS-CoV-2 and No Virus groups in adults and children separately.

**Supplemental Data 3**. Differential expression results directly comparing children and adults with SARS-CoV-2, controlling for viral load.

**Supplemental Data 4**. GSEA applied to the direct comparison between children and adults with SARS-CoV-2.

**Supplemental Data 5**. *In silico* deconvolution of cell type proportions.

## Figures

**Figure 3–figure supplement 1:**
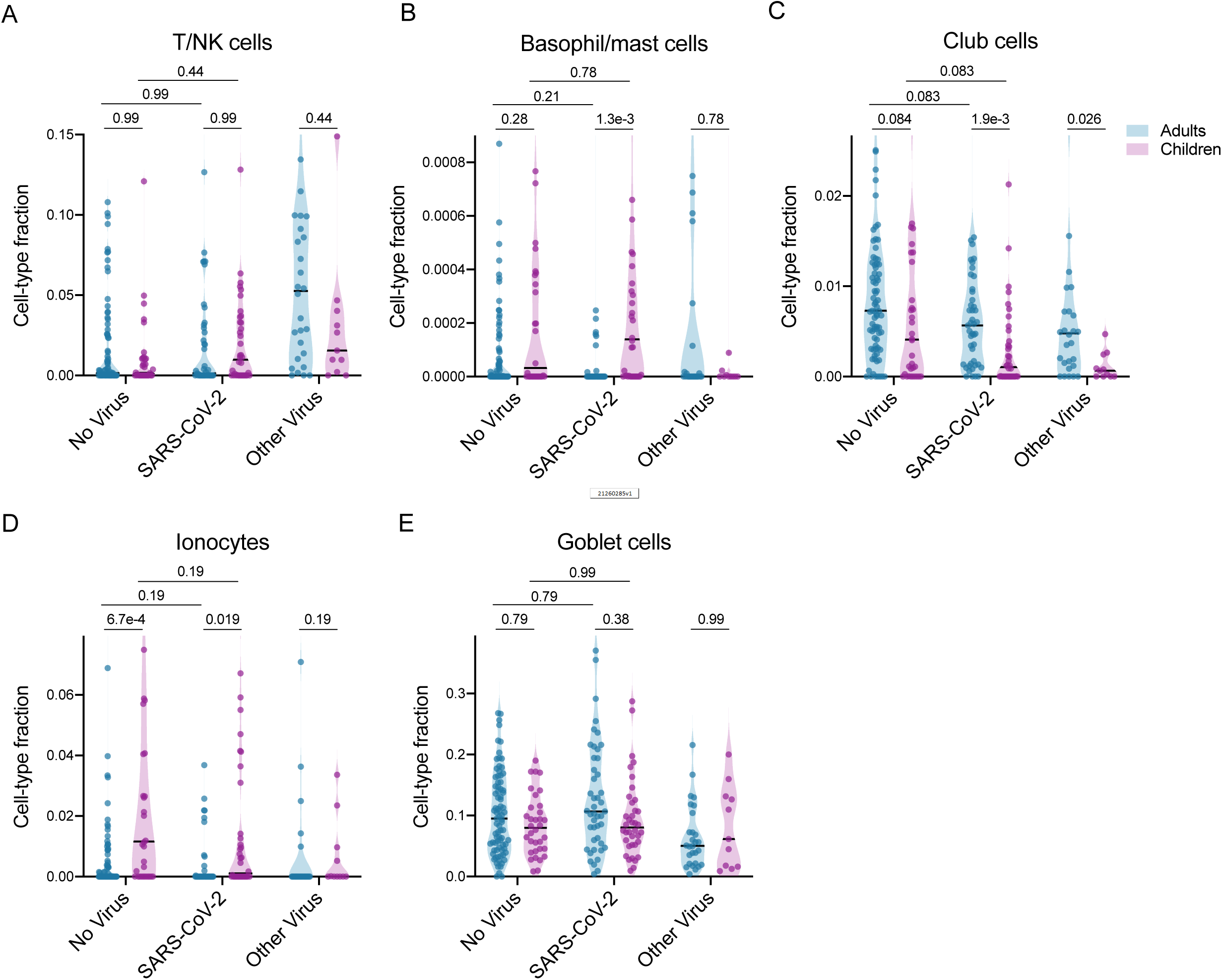
Additional cell-types. Additional cell types included in the cell-type proportions analysis.

## Notes

### Competing Interest Statement

The authors have declared no competing interest.

### Author Declarations

UCSF Specimens and data were obtained under the UCSF Institutional Review Board (IRB) IRB protocol #17-24056, which granted a waiver of consent. CHCO specimens and data were obtained under IRB protocols #0865, #20-1617 and #20-0972, which also granted a waiver of consent.

